# Immune-inflammatory Indices as Predictors of Complete Response to Neoadjuvant Chemoradiotherapy in Locally Advanced Rectal Cancer: A Systematic Review and Meta-Analysis of Observational Studies

**DOI:** 10.64898/2026.01.05.26343418

**Authors:** Mohammadsadra Shamohammadi, Yasaman Gholinezhad, Fereshte Abdolvand, Armaghan Abbasi Garavand, Seyed Hamzeh Mousavie, Mansour Bahrdoust, Marina Yiasemidou

**Author notes:** **Corresponding authors:** Marina Yiasemidou, M.D, MBBS, MSc, PhD, FRCS Consultant colorectal surgeon, United Kingdom Mansour Bahrdoust, PhD, Department of Epidemiology, School of Public Health, Shahid Beheshti University of Medical Sciences, Tehran, Iran. These authors contributed equally as first authors (Mohammadsadra Shamohammadi, Yasaman Gholinezhad).

## Abstract

**Background:** Pathologic complete response (pCR) after neoadjuvant chemoradiotherapy (nCRT) for locally advanced rectal cancer (LARC) is associated with improved survival and organ preservation. Available blood-based inflammatory indices may help to predict pCR and guide individualized treatment.

**Methods:** We conducted a comprehensive search of PubMed, Web of Science, Scopus, and Embase, and Cochrane Library to identify studies including adult patients with pathologically confirmed LARC treated with nCRT followed by total mesorectal excision. We included studies that reported categorical analyses of pre-nCRT NLR (neutrophil-to-lymphocyte ratio), PLR (platelet-to-lymphocyte ratio), or SII (systemic immune-inflammation) in relation to pCR. Random-effects models were used to generate pooled odds ratios [1] for high versus low index values; heterogeneity, small-study effects, meta-regression, and certainty of evidence (GRADE) were assessed.

**Results:** Twenty-four studies (20 NLR, 12 PLR, and 5 SII) met the inclusion criteria; all were rated as high quality according to the Newcastle–Ottawa Scale. Higher pre-nCRT NLR was associated with reduced odds of pCR (OR 0.60, 95% CI 0.49–0.71; I²=35.4%). Elevated PLR similarly predicted lower pCR rates (OR 0.53, 95% CI 0.37–0.69; I²=54.7%), as did higher SII (OR 0.42, 95% CI 0.26–0.58; I²=27.9%). Associations were consistent across cut-off values and geographic regions. Meta-regression suggested that age, stage, sample size, and cut-off selection partially explained between-study heterogeneity. Evidence of small-study effects was observed for NLR and PLR.

**Conclusions:** Higher pre-nCRT NLR, PLR, and SII predict reduced pCR rates in LARC following nCRT. These inexpensive, available indices may refine pre-nCRT risk stratification and selection for organ-preserving.

## Background

Colorectal cancer is classified as the third leading cause of cancer in worldwide and second cause of mortality. Rectal cancer is accounts for one third of colorectal cancer [2] and remains a major challenge due to its growing prevalence and high rates of recurrence and mortality [3, 4]. Locally advanced rectal cancer (LARC) is defined as tumor invasion beyond the muscularis propria and and/or involvement of regional lymph nodes and is generally managed with neoadjuvant chemoradiotherapy (nCRT) followed by total mesorectal excision (TME) and adjuvant chemotherapy [5] or nonoperative “watch-and-wait” management in carefully selected patients [6, 7].

Pathologic complete response (pCR), defined as the absence of residual tumor in the resected rectum and regional lymph nodes after nCRT, is associated with better survival outcomes and a reduced risk of local recurrence [8–10]. However, pCR is occurred around 15 to 20 % of patients, and the rest experience partial response or show resistance to nCRT [11, 12]. The wide variability in treatment response emphasizes the need for reliable predictors of pCR to guide risk-adapted strategies, such as selecting candidates for organ preservation or, alternatively, for treatment intensification to optimize oncological outcomes [13].

Cancer-associated inflammation, a key hallmark of the disease, derives tumorigenesis, progression, and therapeutic response through the dynamic interplay of cellular and molecular components within the tumor microenvironment. The interaction between tumor cells, stromal elements, and immune cells in the tumor microenvironment leads to both local and systemic inflammatory responses, which can be measured using routine blood tests [14, 15]. Neutrophils, which increase during inflammation, act in a pro-tumorigenic manner by promoting tumor initiation, growth, and metastatic spread [16]. Lymphocytes, in contrast, represent the host’s protective anti-tumor immune surveillance, with their activity directly combating tumor progression [17]. Platelets also facilitate tumor growth by aiding cancer cell extravasation and metastatic dissemination [18]. Composite systemic inflammatory indices derived from these cell counts provide a simple, non-invasive measure of the balance between pro-tumor and antitumor forces.

Among these indices, neutrophil-to-lymphocyte ratio (NLR), platelet-to-lymphocyte ratio (PLR) have been widely investigated as prognostic markers across multiple malignancies, including colorectal cancer. These indices are cost-effective and easily derived from routine complete blood counts, offering a practical reflection of the systemic inflammatory status and host-tumor immune equilibrium.

Several studies have suggested that elevated pre-nCRT NLR, PLR, or SII (systemic immune-inflammation) are associated with a lower likelihood of achieving pCR after nCRT in LARC [19–22]. However, other studies have not confirmed these associations [23–25], and differences in sample size, cut-off definitions, and adjustments for confounding factors have led to uncertainty regarding their prognostic value. Given the increasing interest in inflammatory indices and the inconsistent findings across studies, a comprehensive synthesis of the available evidence is necessary. Therefore, we aim to conducted a systematic review and meta-analysis of observational studies to evaluate the relationship between pre-nCRT NLR, PLR, and SII and pCR in patients with LARC undergoing nCRT.

## Methods

### Study Design and Search Strategy

This review was designed and reported in accordance with the Preferred Reporting Items for Systematic Reviews and Meta-Analyses (PRISMA) guidelines [26]. The protocol was prospectively registered in PROSPERO (ID: CRD420251181759). We conducted comprehensive searches of PubMed (MEDLINE), Web of Science, Scopus, Embase, Cochrane Library, and Google Scholar from inception to 15 November 2025. The search was subsequently updated on 31 December 2025 to identify any additional eligible studies published since the initial search. Our search included terms for “neutrophil-to-lymphocyte ratio”, “platelet-to-lymphocyte ratio”, “systemic immune-inflammation”, “pathological complete response”, and “locally advanced rectal cancer” along with related keywords. Search terms were combined using Boolean operators (AND/OR) and included synonyms and associated terms for each concept. The exact strategies for each database appear in supplementary (Table S1). The reference lists of relevant articles were manually screened to identify any other related studies.

### Inclusion and Exclusion Criteria

The inclusion criteria for the study were as follows: (1) All original studies involving adult patients with pathologically confirmed LARC who underwent TME; and (2) studies that evaluated the association between pre-nCRT NLR, PLR, or SII and pCR, and reported effect estimates (odds ratios [ORs] with 95% confidence intervals) comparing high versus low values for these indices or provided sufficient data to calculate them. We excluded studies that: (1) assessed pathological good response rather than pCR as the outcome; (2) reported clinical and/or radiological complete response, or combined these outcomes with pCR in a way that made extraction of pCR-related data impossible; [27] included patients managed with a watch and wait strategy, unless these cases could be clearly separated; (4) collected blood samples for NLR, PLR, or SII only after nCRT; (5) reported outcomes solely using continuous ORs without categorical comparisons; and (6) were case reports, case series, letters to the editor, congress abstracts, publications that were not in English, or articles without accessible full texts. Across all included studies, the indices were calculated using the same formulas: NLR = neutrophils / lymphocytes, PLR = platelets / lymphocytes, and SII = (platelets × neutrophils) / lymphocytes.

### Data Extraction

Two reviewers independently screened all titles and abstracts. A third reviewer evaluated the full texts, and any discrepancies at this stage were resolved by a fourth reviewer. For each included study, two reviewers independently extracted data using a structured data extraction sheet. The following data were extracted: first author, publication year, study design, country, sample size, patient demographics such as gender, age, and, TNM stage, neoadjuvant treatment details, interval between nCRT and surgery, and details of surgical approach when available. We also collected the reported cut-off values for NLR, PLR, and SII, including the methods used to determine each cut-off, and the number of patients above and below the cut-off in both the pCR and non-pCR groups. Effect estimates [1] with 95% confidence intervals were extracted when reported, or calculated from documented data when possible. Any discrepancies in extracted data were resolved by a third reviewer.

### Quality Assessment

Methodological quality was evaluated independently by two reviewers and any discrepancies during quality assessment were resolved by a third reviewer as well. For observational studies, the assessing tool was the Newcastle-Ottawa Scale , which examines bias across three domains: selection of participants, comparability of study groups, and assessment of outcomes [28]. Each observational study got a score up to nine points based on NOS criteria. Studies were classified as high quality (7-9), moderate quality (4-6), or low quality (0-3). The Cochrane Risk of Bias 2 (RoB 2) tool was designated for randomized trials [29]; however, since no RCTs met the inclusion criteria, this tool was not applied.

### Certainty of Assessment

The certainty of the evidence for each outcome was evaluated using the Grading of Recommendations, Assessment, Development and Evaluations (GRADE) approach [30]. Because all included studies were observational, certainty ratings started at ‘low’ and were adjusted based on GRADE domains including risk of bias, inconsistency, indirectness, imprecision, and publication bias. Any discrepancies during quality or certainty assessments were resolved by a third reviewer as well.

### Statistical analysis

Data analysis was performed using Stata, version 17, with a random-effects model. ORs with 95% CIs were calculated to assess the association of NLR, PLR, and SII with pCR. Descriptive results were summarized in tables and figures. Heterogeneity across studies was evaluated using the Q test and the I² statistic, with I² values interpreted as follows: 0–40% as little or no heterogeneity, 30–60% as moderate heterogeneity, 50–90% as substantial heterogeneity, and 75–100% as considerable heterogeneity. Egger’s test was used to assess publication bias, and the results were displayed in funnel plots; when publication bias was identified, trim-and-fill analysis was applied. Subgroup analyses were conducted according to geographic region and cut-off values for NLR, PLR, and SII. Sensitivity analyses were used to examine the impact of individual studies on the association between these indices and pCR. Meta-regression was performed to investigate potential sources of between-study heterogeneity.

## Results

### Study selection

A total of 206 articles were identified through database searches (PubMed n = 88, Embase n = 187, Scopus n = 211, Web of Science n = 84, Cochrane Library n = 5). After removing 281 duplicates, 294 articles were screened based on their titles and abstracts. A total of 235 articles were excluded. Fifty-nine articles underwent full-text screening, and 35 were excluded as they did not meet the selection criteria. Ultimately, 24 studies were included in this meta-analysis (Figure 1).

**Figure 1.**
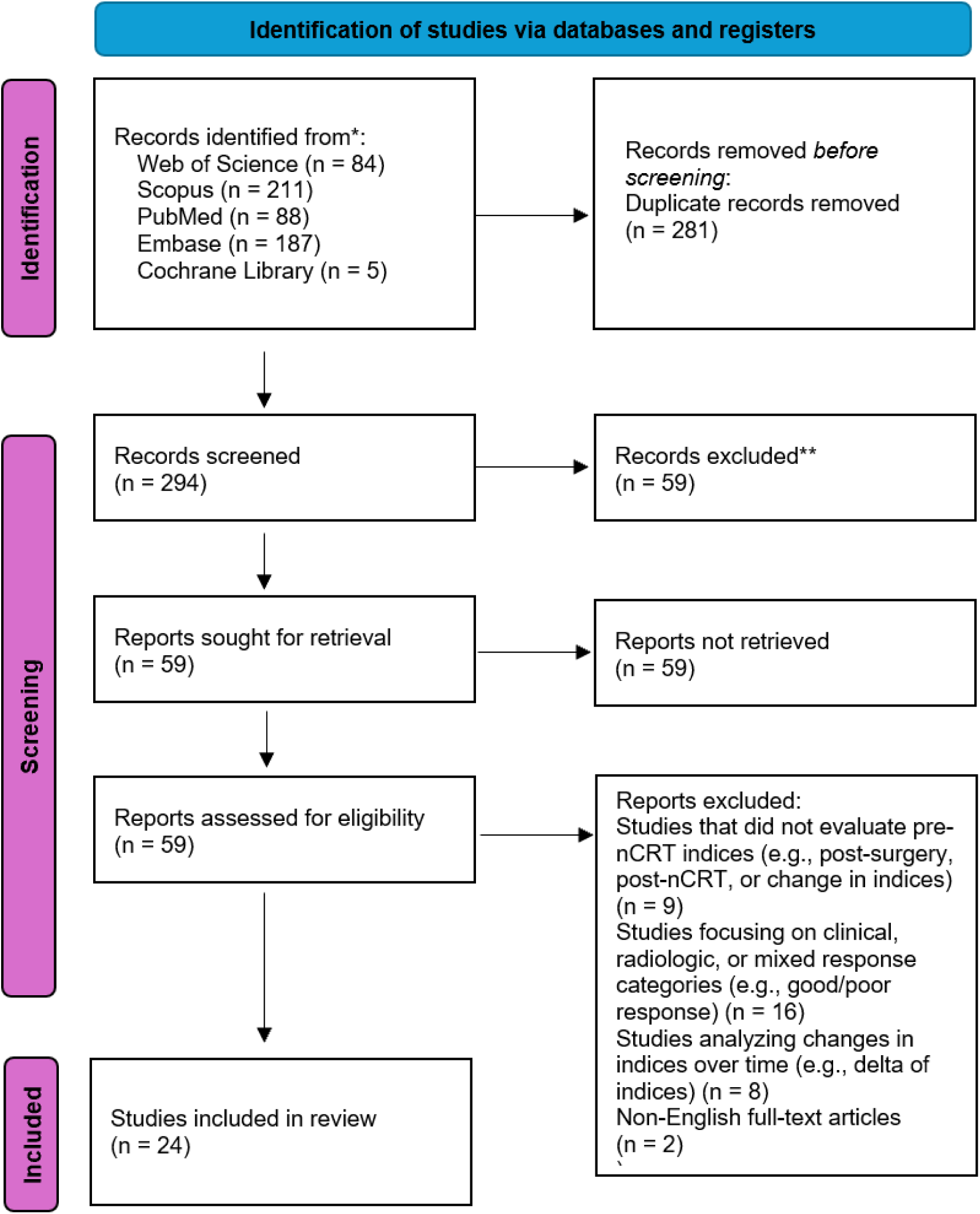
PRISMA flow diagram

### Study characteristics

The main characteristics of the included studies are summarized in Table 1. A total of 24 studies [19–25, 31–47], published between 2013 and 2025, from 9 countries across Asia, Europe, and the Americas were included. All studies were retrospective cohort designs, evaluating adult patients with pathologically confirmed LARC treated with nCRT followed by TME. The sample sizes ranged from 56 to 1237 patients, with a total of 13,369 participants. The male-to-female ratio across studies was approximately 1.9:1, with 6,579 male patients and 3,469 female patients.

**Table 1.**
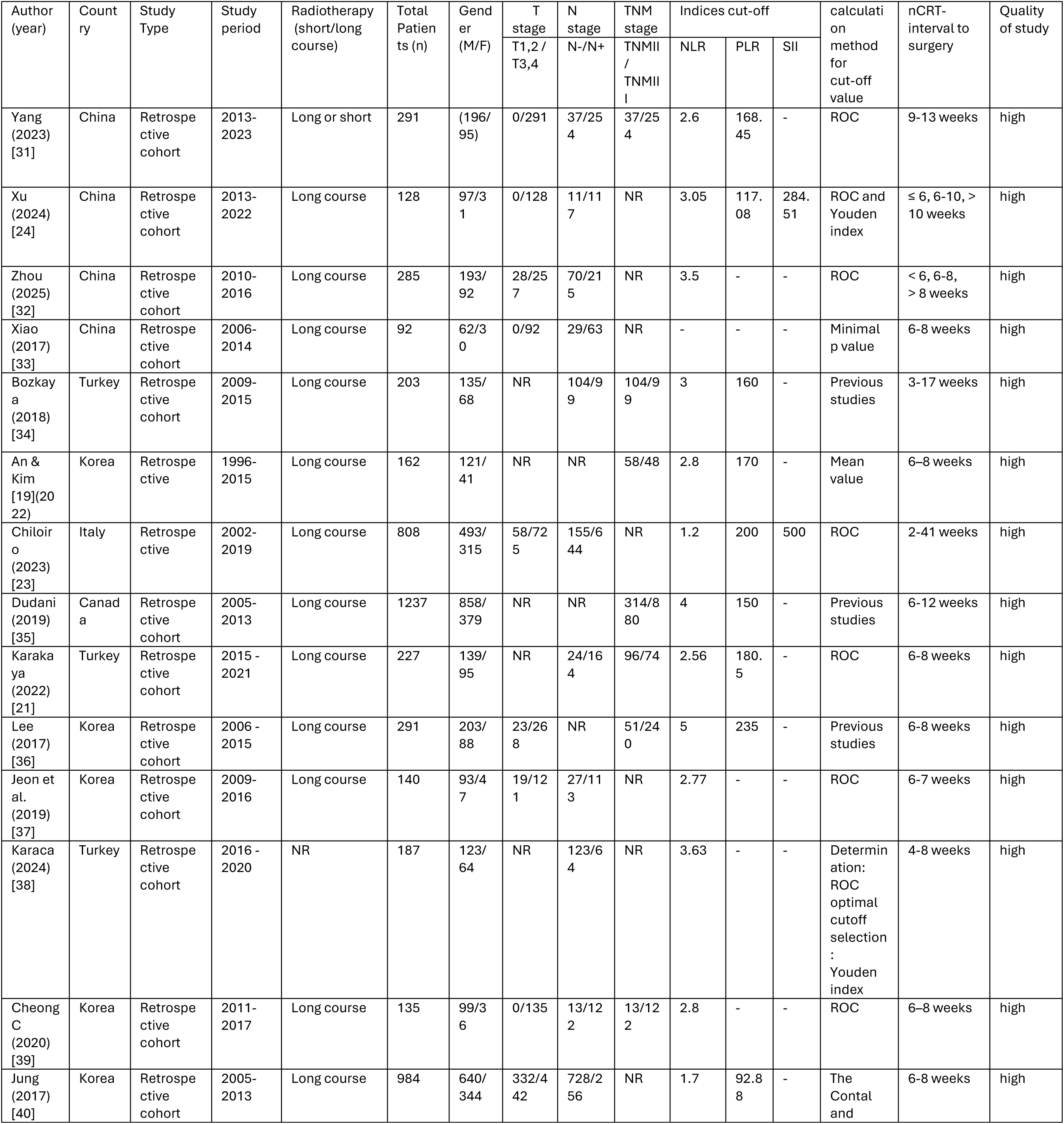
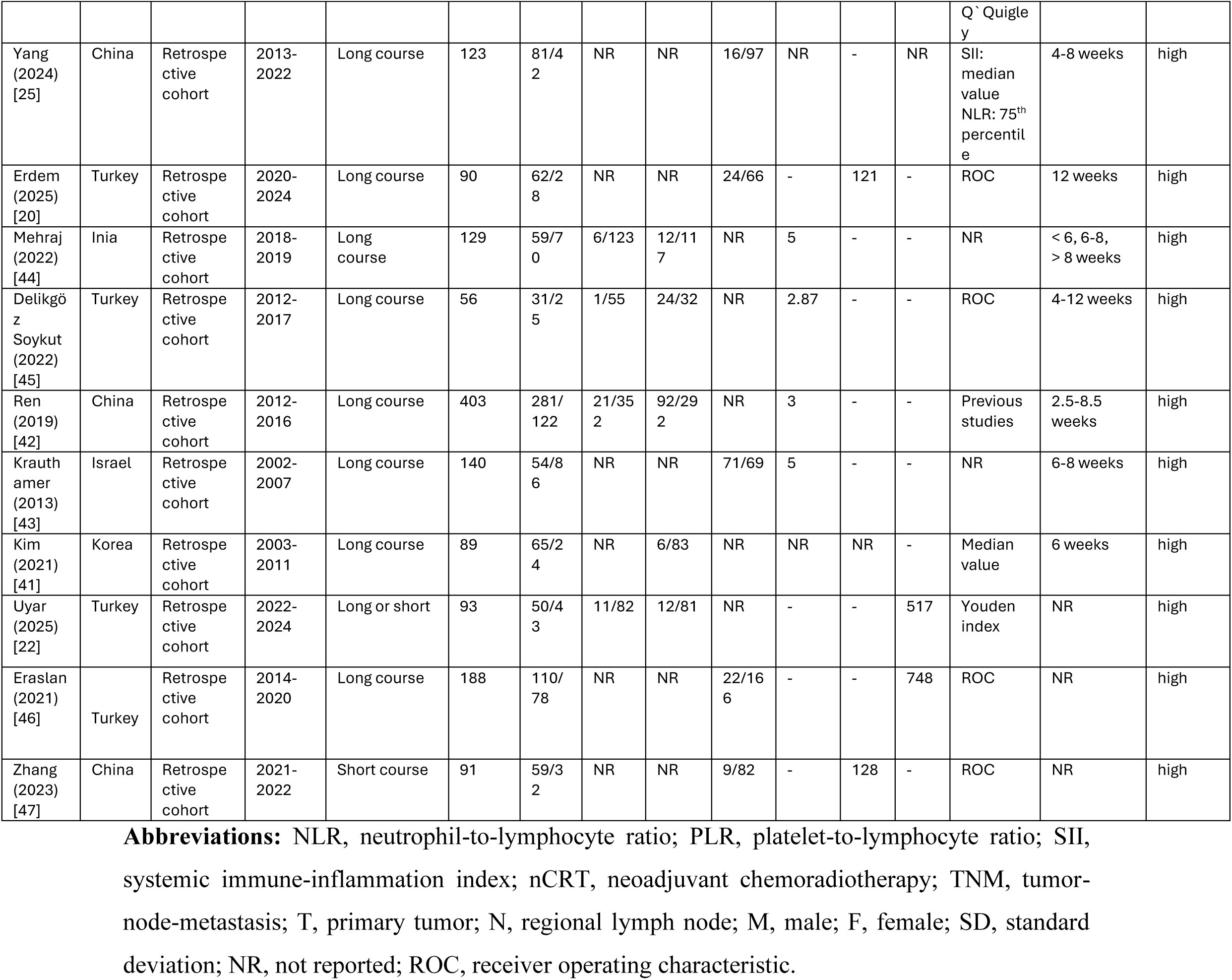
Baseline Characteristics

**Table 2.** Subgroup analyses of the association between pretreatment immune-inflammatory indices and pCR after nCRT in LARC.

| **NLR** | Subgroup factor | Pooled OR (95% CI) | I<sup>2</sup> (%) | p-value for heterogeneity |
| --- | --- | --- | --- | --- |
| Cut-off value | < 3 | 0.57 (0.45–0.69) | 0.0 | 0.523 |
| Cut-off value | ≥ 3 | 0.67 (0.46–0.87) | 57.5 | 0.012 |
| Region | Asia | 0.60 (0.46–0.73) | 44.3 | 0.038 |
| Region | Europe | 0.62 (0.41–0.82) | 14.9 | 0.319 |
| Region | Americas | 0.78 (0.27–1.29) | – | – |
| **PLR** |  |  |  |  |
| Cut-off value | ≤ 150 | 0.59 (0.25–0.94) | 69.7 | 0.010 |
| Cut-off value | > 150 | 0.54 (0.35–0.72) | 42.1 | 0.110 |
| Region | Asia | 0.59 (0.32–0.85) | 64.2 | 0.010 |
| Region | Europe | 0.51 (0.30–0.72) | 43.8 | 0.130 |
| **SII** |  |  |  |  |
| Cut-off value | < 500 | 0.47 (0.23–0.71) | 6.2 | 0.302 |
| Cut-off value | ≥ 500 | 0.42 (0.17–0.67) | 48.5 | 0.143 |

**Abbreviations:** p, p-value; NLR, neutrophil-to-lymphocyte ratio; PLR, platelet-to-lymphocyte ratio; TNM, tumor–node–metastasis; pre-nCRT, pretreatment neoadjuvant chemoradiotherapy.

Among patients with available staging data, 490 had cT1–2 disease, and 3132 had cT3–4 disease. The total number of patients with N- (node-negative) disease was 1121, and with N+ (node-positive) disease was 2950. Additionally, 785 patients had TNM stage II disease, and 2177 had TNM stage III disease. Most studies used long-course radiotherapy, with the interval from completion of nCRT to surgery ranging from 2 to 41 weeks. Cut-off values for these indices varied across studies, with most being derived from receiver-operating characteristic curve analysis; detailed thresholds are presented in Table 1. All studies were considered to be of high methodological quality according to the NOS, and full NOS scores for each study are reported in Supplementary Table S2.

### NLR predicting pCR

Across 20 studies [19, 21, 23–25, 31–45], a higher pre-nCRT NLR was associated with lower odds of achieving pCR (pooled OR 0.60, 95% CI: 0.49–0.71; I² = 35.4%, p = 0.060). Study-level effects were directionally consistent and no single study dominated the weights (Figure 2).

**Figure 2:**
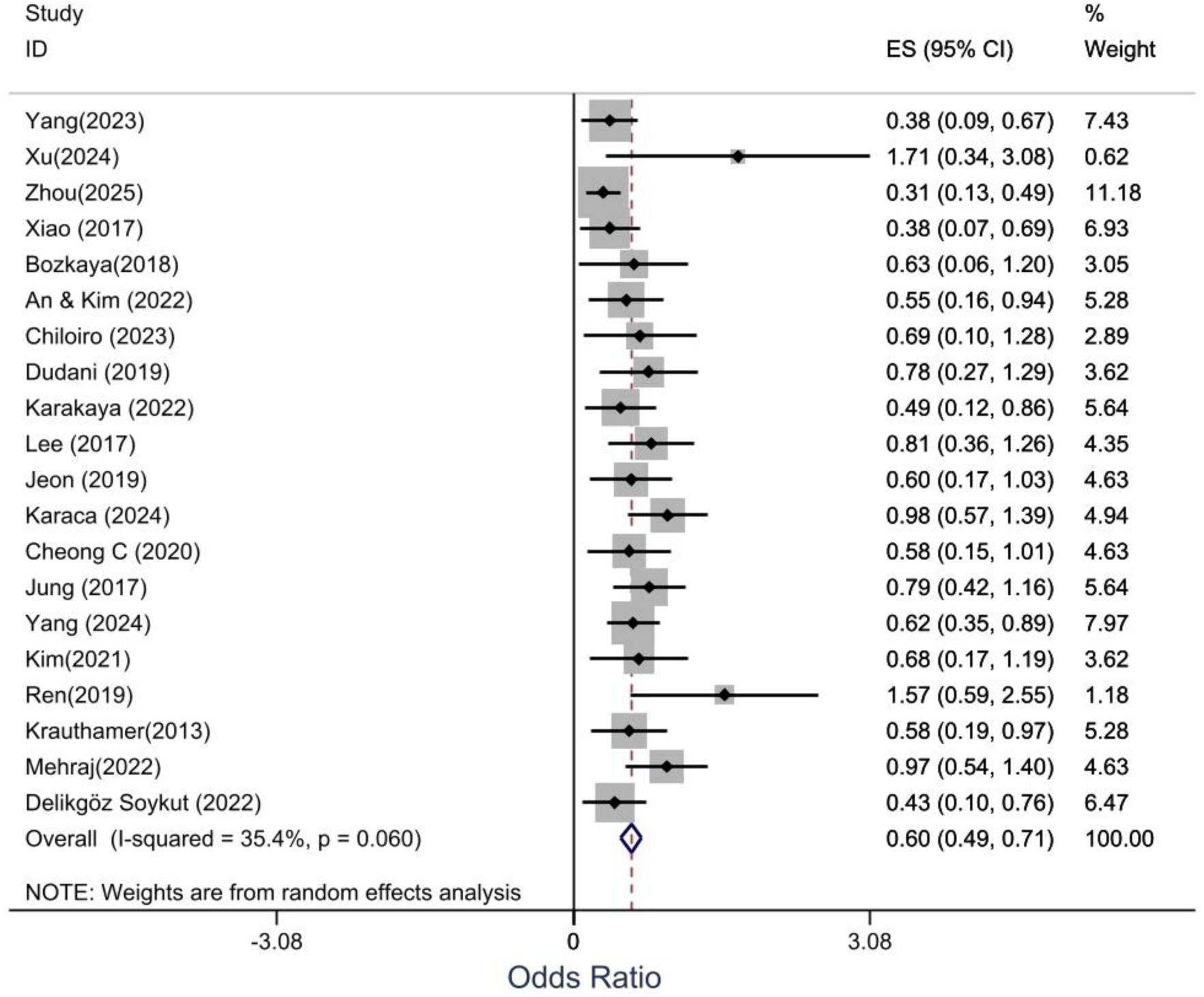
Random–effects meta–analysis of high vs low pre-nCRT NLR and pCR after neoadjuvant chemoradiotherapy in LARC.

### PLR predicting pCR

Across 12 studies [19–21, 23, 24, 31, 34–36, 40, 41, 47], a higher pre-nCRT PLR was associated with lower odds of achieving pCR (pooled OR 0.53, 95% CI 0.37–0.69; I² = 54.7%, p = 0.012), with substantial heterogeneity (I² =54.7%) (Figure 3).

**Figure 3:**
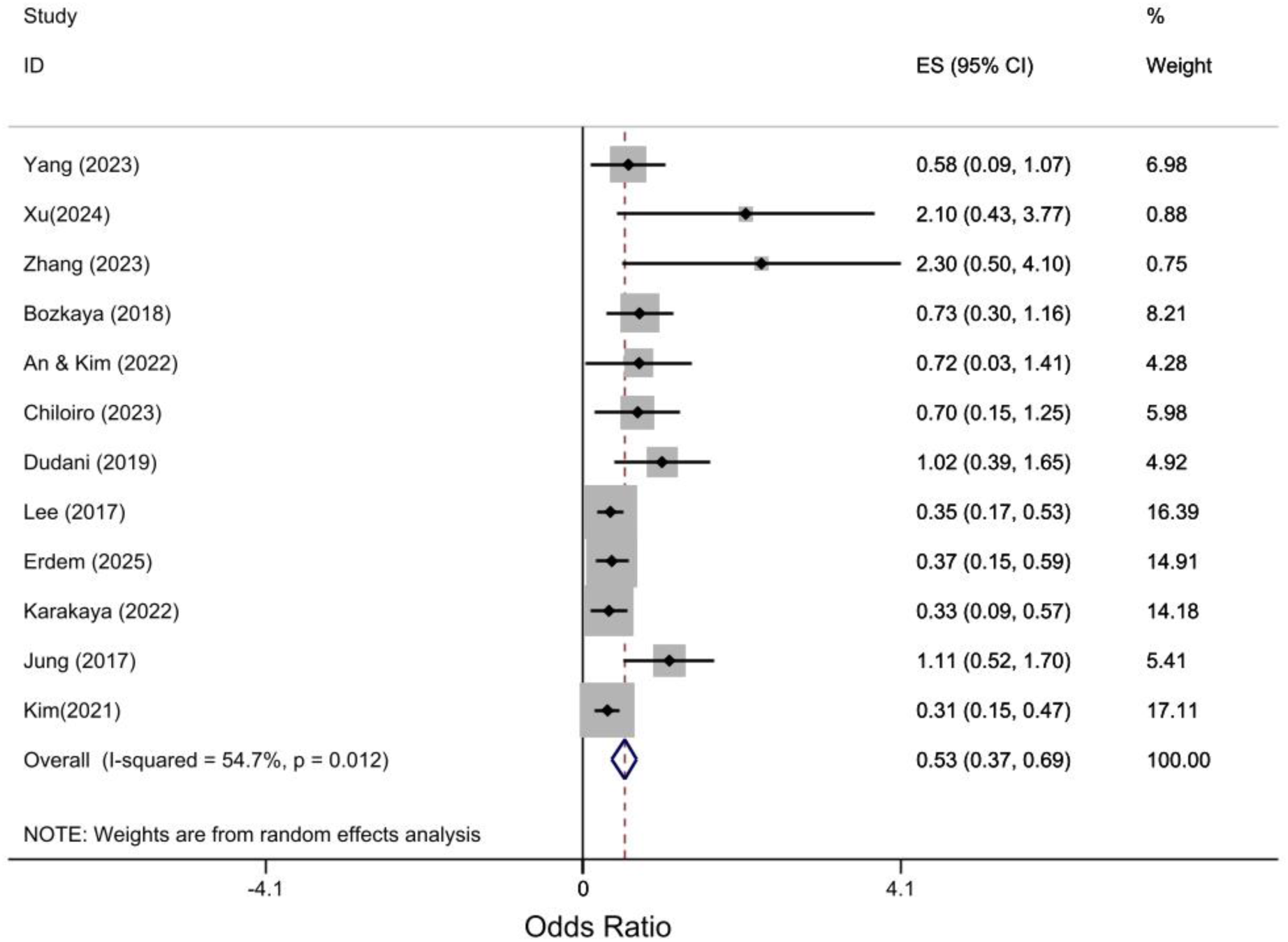
Random-effects Forest plot of high vs low pre-nCRT PLR and pCR.

**Figure 4:**
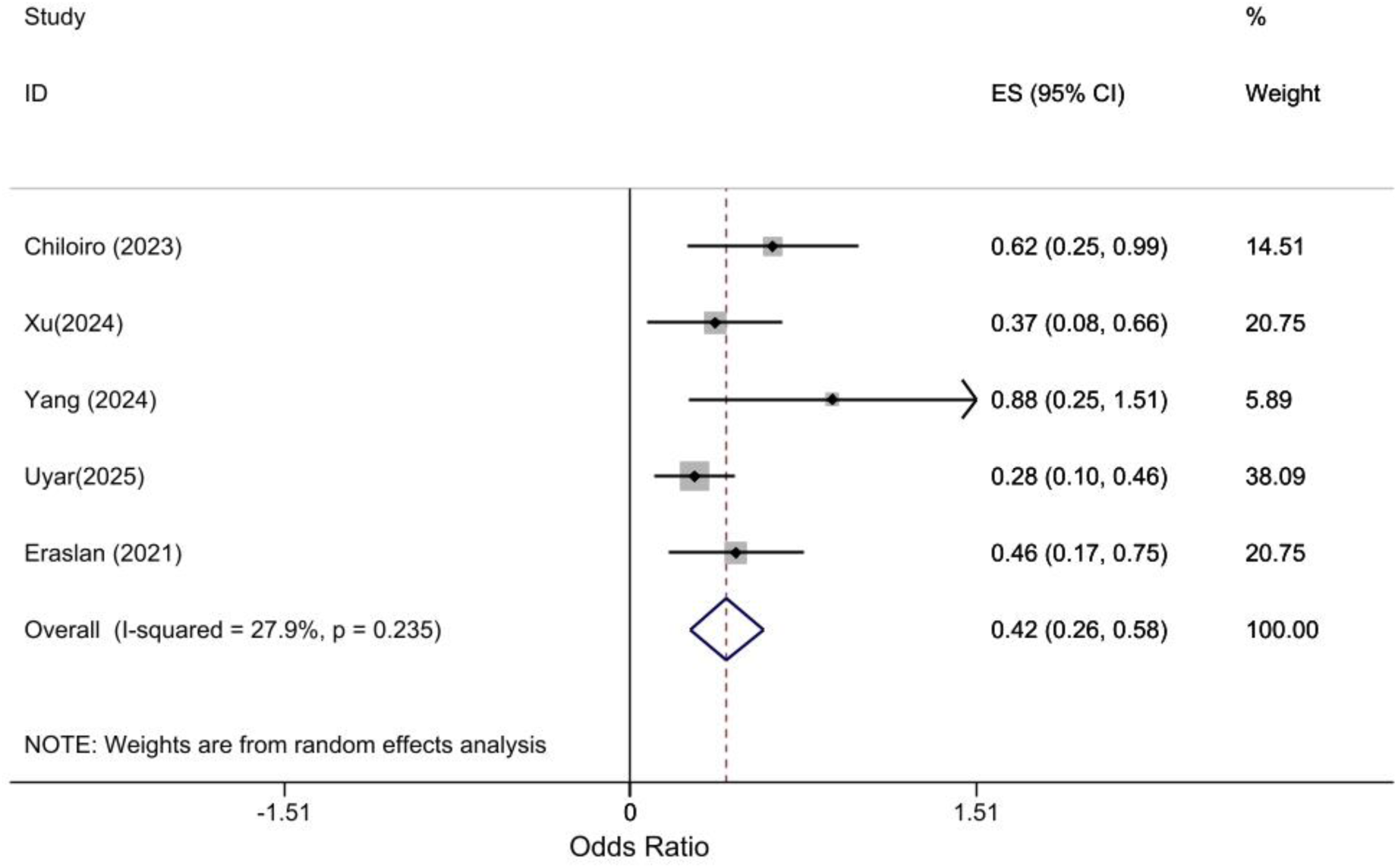
Random–effects meta–analysis of high vs low pre-nCRT SII and pCR after neoadjuvant chemoradiotherapy in LARC.

### SII predicting pCR

Across 5 studies [22–25, 46], a higher pre-nCRT SII was associated with lower odds of achieving pCR (pooled OR 0.42, 95% CI 0.26–0.58). Heterogeneity was low (I² = 27.9%, p = 0.235) (Figure 4).

**Figure 4:**
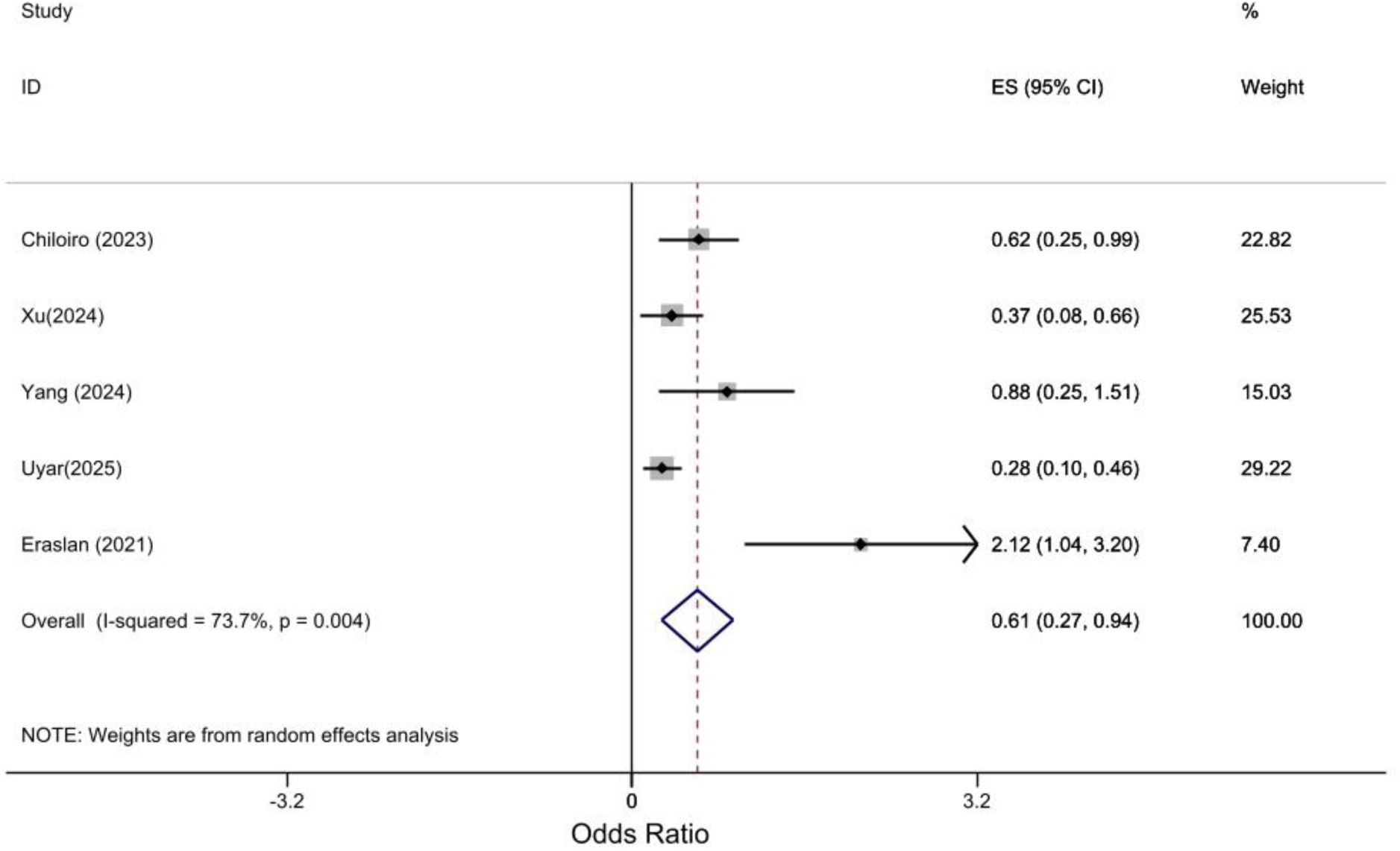
Random-effects Forest plot of high vs low pre-nCRT SII and pCR.

### Subgroup analyses

Subgroup results for NLR, PLR, and SII are summarized in Supplementary Table S2 and illustrated in the corresponding forest plots (Supplementary Figures S1–S3).

For NLR, pooled ORs for studies using thresholds <3 and >3 were 0.57 (95% CI 0.45–0.69) and 0.67 (95% CI 0.46–0.87), respectively. The association was further examined by region, with pooled ORs of 0.60 (95% CI 0.46–0.73) for Asia, 0.62 (95% CI 0.41–0.82) for Europe, and 0.78 (95% CI 0.27–1.29) for the Americas (Supplementary Figure S1). For PLR, studies with thresholds >150 showed a pooled OR of 0.54 (95% CI 0.35–0.72), while studies using thresholds <150 yielded an OR of 0.59 (95% CI 0.25–0.94). By region, pooled ORs were 0.59 (95% CI 0.32–0.85) for Asia and 0.51 (95% CI 0.30–0.72) for Europe (Supplementary Figure S2). For SII, the pooled ORs for studies with sample sizes <500 and >500 were 0.47 (95% CI 0.23–0.71) and 0.42 (95% CI 0.17–0.67), respectively (Supplementary Figure S3).

### Sensitivity analyses

Sensitivity analysis was performed to assess the effect of individual studies on the overall outcome. The results of the sensitivity analysis showed that studies with a sample size (<500 patients) had the highest effect on the association between NLR with pCR (Supplementary Figure S4) and PCR and pCR (Supplementary Figure S5). Studies with a sample size (<100 patients) also had the highest effect on the association between SSI and pCR (Supplementary Figure S6).

### Publication Bias

Egger’s test provided evidence of publication bias for the association of NLR (t = 2.6; 95% CI 1.10–3.11; p = 0.021) and PLR (t = 1.8; 95% CI 0.51–3.25; p = 0.027) with complete response, which was in line with the asymmetry observed in the corresponding funnel plots (Figure 5A, B). Given the presence of publication bias, the trim-and-fill method was used. This analysis indicated that eight studies were potentially missing due to publication bias, and the pooled effects were adjusted accordingly (Supplementary Figure S3A, B). In contrast, Egger’s test did not demonstrate evidence of publication bias for SII (t = 2.9; 95% CI −0.53–6.41; p = 0.058), and the distribution of studies in the funnel plot appeared approximately symmetric (Figure 5C).

**Figure 5:**
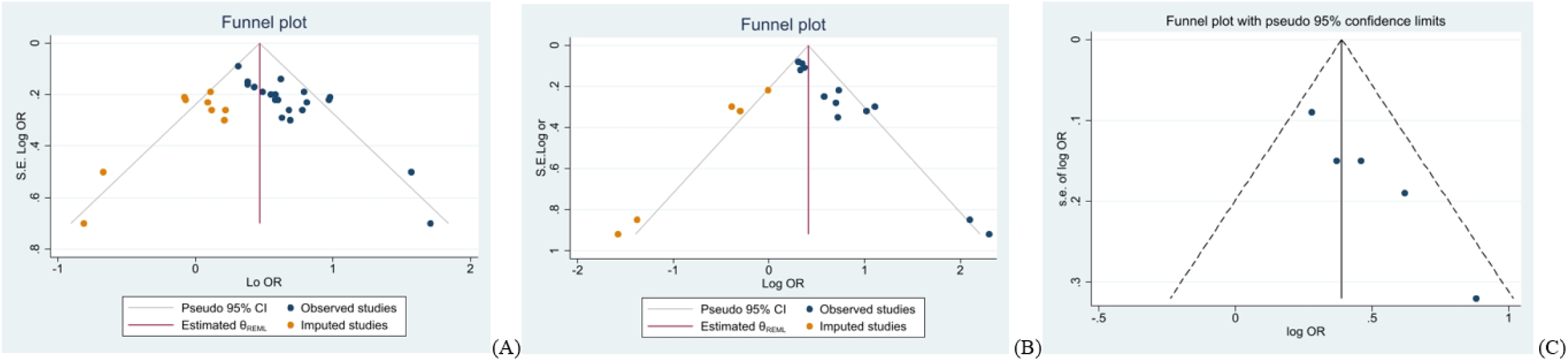
Bias publication assessment in the funnel plot for NLR (A), PLR rate (B), and SII (C)

### Meta-regression

Meta-regression analysis showed that the age, TNM stage, gender, Pre-nCRT-NLR Cutoff value, quality of studies, geographical regions, and sample size of included studies were associated with heterogeneity between studies to correlate NLR with complete response to treatment. Also, the age, TNM stage, Pre-nCRT-PLR Cutoff value, geographical regions, and sample size of included studies were associated with heterogeneity between studies to correlate PLR with complete response to treatment (Table 3).

**Table 3.** Meta-regression model of the effect of variables on effect size

| Variable | Coefficient<br>$\beta$ | SE | P |
| --- | --- | --- | --- |
| **NLR** |  |  |  |
| Mean Age (Year) | 0.13 | 0.04 | 0.011 |
| TNM stage≥II | 1.08 | 0.12 | 0.001 |
| Sample size | 0.60 | 0.12 | 0.001 |
| Geographical region | -0.19 | 0.1 | 0.044 |
| Pre-nCRT-NLR Cutoff value | 0.27 | 0.08 | 0.001 |
| **PLR** |  |  |  |
| Mean Age (Year) | 0.13 | 0.05 | 0.001 |
| TNM stage≥II | 1.11 | 0.16 | 0.001 |
| Sex (Male) | 0.29 | 0.11 | 0.011 |
| Sample size | 0.65 | 0.17 | 0.004 |
| Geographical region | -0.15 | 0.08 | 0.048 |
| Pre-nCRT-PLR Cutoff value | 0.35 | 0.1 | 0.001 |

**Abbreviations:** $\beta$ , regression coefficient from the meta-regression model; SE, standard error; p, p-value; NLR, neutrophil-to-lymphocyte ratio; PLR, platelet-to-lymphocyte ratio; TNM, tumor–node–metastasis; pre-nCRT, pretreatment neoadjuvant chemoradiotherapy.

### Certainty of evidence (GRADE)

The certainty of evidence for NLR was rated as moderate, while for PLR and SII, it was rated as low. Full domain-level GRADE judgments are available in Supplementary Table S3

## Discussion

This systematic review and meta-analysis synthesized evidence from 24 observational studies to evaluate the prognostic value of pretreatment immune-inflammatory indices NLR, PLR and SII in predicting pCR in patients with LARC who underwent nCRT. Our meta-analysis showed that elevated pretreatment levels of NLR, PLR and SII were associated with reduced odds of achieving pCR, with low to moderate heterogeneity across indices. Meta-regression identified that the age, geographical regions, sample size, TNM stage and pre-nCRT cut-off value are the common sources of variability in NLR and PLR groups. The predictive signal was notable across studies from Asia and Europe.

The results of our study are aligned with previous evidence. Our finding that elevated pre-nCRT inflammatory indices predicts reduced pCR odds (NLR= OR:0.6, PLR=OR:0.53, SII= OR: 0.42) in 13,369 patients aligns with individual rectal cancer studies. Zhou et al. reported that NLR (OR: 0.19) predicted a lower pCR in 285 patients with LARC, confirming directional consistency despite methodological differences [48]. Contrasting our findings, the study by Xu et al. identified an elevated lymphocyte-to-monocyte ratio (LMR > 2.73) as an independent predictor of pCR (OR :4.761), while indices of systemic inflammation (NLR, PLR, SII) showed no significant association in their cohort [24]. Across comparable rectal cancer cohorts is found elevated inflammatory indices predict poor nCRT response, validating our findings [35, 49, 50].

This pattern extends to other malignancies. Breast cancer meta-analysis by Zhao et al., including 7,557 patients reported a negative association between pre-treatment PLR and pCR (HR = 1.51; 95% CI: 1.24-1.84) in patients with breast cancer [51]. Also, in a study by Cullinane et al. in 2020 similarly found that a lower NLR predicts higher pCR rates (OR:1.83, 95% CI 1.15–2.91) in breast cancer [52]. Additionally, the prognostic significance of an elevated pre-treatment NLR, demonstrated by its association with poorer survival and reduced pCR in esophageal cancer (OR: 0.67), supports our parallel finding in rectal cancer [53].

It should be noted that, beyond the classic systemic inflammatory indices such as NLR, PLR, and SII, the results of our meta-analysis are part of a broader landscape of hematologic and nutritional biomarkers. For instance, in breast cancer, studies have validated the Prognostic Nutritional Index (PNI), a marker reflecting both nutritional status and immune competence, as an independent predictor of pCR [54]. Similarly, the serum Albumin-to-Alkaline Phosphatase Ratio (AAPR), an indicator of systemic inflammation and hepatic function, has also demonstrated independent predictive value for pCR in breast cancer [55]. Therefore, integrating diverse, low-cost laboratory indices into multivariable models is advisable to improve pre-treatment risk stratification in oncology.

To maintain analytical uniformity, our inclusion criteria were restricted to studies that employed a categorical approach based on optimal cut-off points determined by ROC curve analysis for NLR:1.2-5.0, PLR:117-235, SII:284-797, enabling clinical risk stratification despite cut-off heterogeneity [56]. In contrast, Shi et al. (2022) analyzed continuous pretreatment NLR in 59 LARC patients, identifying raw NLR as an independent pCR predictor (OR: 4.205 per unit increase, P=0.027) with lower values in pCR. Similarly, articles analyzed inflammatory indices as continuous variables, confirming the same directional relationship with pCR without binary categorization [57, 58].

The inverse relationship observed for all three indices aligns with the pattern of cancer-associated inflammation. Mechanistically, high neutrophils that lead to producing VEGF, FGF [59, 60] which released via MMP-9 degradation of the extracellular matrix, directly driving tumor angiogenesis [61]. Also, high levels of IL-10, TGF-β are associated with neutrophilia which promotes angiogenesis and immunosuppression [62, 63]. These cytokines suppress cytotoxic CD8+ T-cells and recruit regulatory T-cells, creating an immunosuppressive niche [64]. High platelet counts drive metastasis [65]. Platelets secrete TGF-β, which activates the TGF-β/Smad pathway in tumor cells, inducing Epithelial-Mesenchymal Transition (EMT). This process allows tumor cells to detach and metastasize [66, 67]. Taken together, these mechanisms support our results that the pre-treatment elevation of NLR, PLR, and SII quantifies a systemic pro-tumorigenic shift that compromises the efficacy of chemoradiotherapy, thereby reducing the likelihood of a pCR.

The results of this meta-analysis should be interpreted cautiously due to several limitations inherent to the observational study designs. In particular, there was heterogeneity in the treatment regimens, including conventional nCRT versus total neoadjuvant therapy and different chemotherapy backbones, in the cut-off values and timing of blood sampling used to define high versus low indices, and in the level of statistical adjustment. Several studies reported only unadjusted ORs, which raises the possibility of residual confounding. In addition, we observed evidence of small-study effects and possible publication bias for NLR and PLR, which means that the true magnitude of association may be smaller than our pooled estimates. For SII, only five studies were available, so the results for this index should be regarded as less robust than those for NLR and PLR.

Despite the study’s limitations, the findings highlighted the role of pre-treatment systemic inflammatory indices as practical, inexpensive, and universally available prognostic biomarkers in LARC. Incorporating these indices into clinical decision-making frameworks may aid risk stratification by identifying patients with a lower likelihood of achieving pCR, which helps set realistic expectations and could guide different discussions about treatment strategies in clinical trials. Furthermore, these biomarkers may enable more personalized therapy by selecting patients with a high inflammatory burden for novel approaches, such as combining anti-inflammatory or immunomodulatory agents with standard nCRT [68]. While our meta-analysis identified elevated baseline inflammatory indices as biomarkers associated with a lower probability of pCR following standard chemoradiotherapy, there is evidence suggesting therapeutic strategy to overcome such resistance, demonstrating that integrating immunotherapy with total neoadjuvant therapy (TNT) can achieve substantially higher pCR rates in pMMR/MSS LARC and guide patients to Watch-and-Wait strategy for patients [69].

However, translation of these findings into clinical practice requires overcoming several obstacles, including validating optimal standardized cut-off values through large, prospective studies and evaluating dynamic changes in these indices during and after nCRT for improved prediction. Also, integrating inflammatory markers with other prognostic factors, such as MRI-based regression grade, CEA, and molecular biomarkers, into comprehensive predictive models is warranted. Interestingly, due the inverse relation between indices and treatment response, investigating the mechanistic relation between systemic inflammation and radiotherapy resistance can suggest future novel therapeutic targets. These findings collectively suggest that pretreatment NLR, PLR, and SII are promising and easily accessible prognostic markers for response to nCRT in LARC. However, they emphasize the necessity for prospective, standardized validation before these markers can be routinely used in clinical practice.

## Conclusion

Results of the current study demonstrate that higher pre-nCRT NLR, PLR, and SII are associated with a lower likelihood of achieving pCR in patients with LARC and may provide additional information for pre-treatment risk stratification in this setting. These immune-inflammatory indices are inexpensive and routinely available from complete blood counts, making them attractive candidates for incorporation into nomograms and other prediction tools. Prospective studies with standardized cut-off values and predefined adjustment for confounders are needed to confirm these findings and to establish the definitive role of immune-inflammatory indices in prognostic models for predicting pCR and guiding the management of LARC.

## Abbreviations

CI: confidence interval
CRT: chemoradiotherapy
GRADE: Grading of Recommendations, Assessment, Development and Evaluation
LARC: locally advanced rectal cancer
nCRT: neoadjuvant chemoradiotherapy
NLR: neutrophil-to-lymphocyte ratio
NOS: Newcastle-Ottawa Scale
NR: not reported
OR: odds ratio
pCR: pathologic complete response
PLR: platelet-to-lymphocyte ratio
PRISMA: Preferred Reporting Items for Systematic Reviews and Meta-Analyses
PROSPERO: International Prospective Register of Systematic Reviews
RCT: randomized controlled trial
RoB 2: Risk of Bias 2 tool
ROC: receiver operating characteristic
SD: standard deviation
SE: standard error
SII: systemic immune-inflammation index
TME: total mesorectal excision
TNM: tumor-node-metastasis staging system

## Data Availability

All data produced in the present work are contained in the manuscript.

## Acknowledgements

NA.

## Data availability

The datasets generated or analyzed during the current study are available from the corresponding author upon reasonable request.

## Author contributions (CRediT)

Conceptualization: M.S and Y.G; Methodology: M.S and M.Y; Formal analysis: M.B and A.A.G; Data curation: Y.G, F.A, and A.A.G; Investigation: M.S, M.Y; Validation: M.B and S.H.M; Writing – original draft: M.S, Y.G, and F.A; Writing – review and editing: M.S, M.Y, S.H.M, and M.B; Visualization: M.S, M.B; Supervision: M.B, S.H.M; Project administration: M.S, Y.G.

## Declarations

### Human Ethics and Consent to Participate declarations

Not applicable.

### Ethics approval and consent to participate

Not applicable. This study is a systematic review and meta-analysis of published, de-identified data and did not involve direct contact with human participants.

## Consent to participate

Not applicable.

## Consent for publication

Not applicable.

## Protocol registration

PROSPERO CRD420251181759.

## References

1. Morson BC: Carcinoma arising from areas of intestinal metaplasia in the gastric mucosa. British Journal of Cancer 1955, 9(3):377–385.

2. Bray F, Laversanne M, Sung H, Ferlay J, Siegel RL, Soerjomataram I, Jemal A: Global cancer statistics 2022: GLOBOCAN estimates of incidence and mortality worldwide for 36 cancers in 185 countries. CA Cancer J Clin 2024, 74(3):229–263.

3. Fadlallah H, El Masri J, Fakhereddine H, Youssef J, Chemaly C, Doughan S, Abou-Kheir W: Colorectal cancer: Recent advances in management and treatment. World J Clin Oncol 2024, 15(9):1136–1156.

4. Lefèvre JH, Mineur L, Cachanado M, Denost Q, Rouanet P, de Chaisemartin C, Meunier B, Mehrdad J, Cotte E, Desrame J et al: Does A Longer Waiting Period After Neoadjuvant Radio-chemotherapy Improve the Oncological Prognosis of Rectal Cancer?: Three Years’ Follow-up Results of the Greccar-6 Randomized Multicenter Trial. Ann Surg 2019, 270(5):747–754.

5. Ali F, Keshinro A, Weiser MR: Advances in the treatment of locally advanced rectal cancer. Ann Gastroenterol Surg 2021, 5(1):32–38.

6. Scott AJ, Kennedy EB, Berlin J, Brown G, Chalabi M, Cho MT, Cusnir M, Dorth J, George M, Kachnic LA et al: Management of Locally Advanced Rectal Cancer: ASCO Guideline. J Clin Oncol 2024, 42(28):3355–3375.

7. Hofheinz RD, Fokas E, Benhaim L, Price TJ, Arnold D, Beets-Tan R, Guren MG, Hospers GAP, Lonardi S, Nagtegaal ID et al: Localised rectal cancer: ESMO Clinical Practice Guideline for diagnosis, treatment and follow-up. Ann Oncol 2025, 36(9):1007–1024.

8. Kapiteijn E, Marijnen CA, Nagtegaal ID, Putter H, Steup WH, Wiggers T, Rutten HJ, Pahlman L, Glimelius B, van Krieken JH et al: Preoperative radiotherapy combined with total mesorectal excision for resectable rectal cancer. N Engl J Med 2001, 345(9):638–646.

9. Conroy T, Bosset JF, Etienne PL, Rio E, François É, Mesgouez-Nebout N, Vendrely V, Artignan X, Bouché O, Gargot D et al: Neoadjuvant chemotherapy with FOLFIRINOX and preoperative chemoradiotherapy for patients with locally advanced rectal cancer (UNICANCER-PRODIGE 23): a multicentre, randomised, open-label, phase 3 trial. Lancet Oncol 2021, 22(5):702–715.

10. Alexandrescu ST, Dumitru AV, Babiuc RD, Costea RV: Assessment of clinical and pathological complete response after neoadjuvant chemoradiotherapy in rectal adenocarcinoma and its therapeutic implications. Rom J Morphol Embryol 2021, 62(2):411–425.

11. Ryan JE, Warrier SK, Lynch AC, Ramsay RG, Phillips WA, Heriot AG: Predicting pathological complete response to neoadjuvant chemoradiotherapy in locally advanced rectal cancer: a systematic review. Colorectal Dis 2016, 18(3):234–246.

12. Maas M, Nelemans PJ, Valentini V, Das P, Rödel C, Kuo LJ, Calvo FA, García-Aguilar J, Glynne-Jones R, Haustermans K et al: Long-term outcome in patients with a pathological complete response after chemoradiation for rectal cancer: a pooled analysis of individual patient data. Lancet Oncol 2010, 11(9):835–844.

13. Qing S, Gu L, Du T, Yin X, Zhang KJ, Zhang HJ: A Predictive Model to Evaluate Pathologic Complete Response in Rectal Adenocarcinoma. Technol Cancer Res Treat 2023, 22:15330338231202893.

14. Nishida A, Andoh A: The Role of Inflammation in Cancer: Mechanisms of Tumor Initiation, Progression, and Metastasis. Cells 2025, 14(7).

15. de Visser KE, Joyce JA: The evolving tumor microenvironment: From cancer initiation to metastatic outgrowth. Cancer Cell 2023, 41(3):374–403.

16. Khan U, Chowdhury S, Billah MM, Islam KMD, Thorlacius H, Rahman M: Neutrophil Extracellular Traps in Colorectal Cancer Progression and Metastasis. Int J Mol Sci 2021, 22(14).

17. Bai Z, Zhou Y, Ye Z, Xiong J, Lan H, Wang F: Tumor-Infiltrating Lymphocytes in Colorectal Cancer: The Fundamental Indication and Application on Immunotherapy. Front Immunol 2021, 12:808964.

18. Lazar S, Goldfinger LE: Platelets and extracellular vesicles and their cross talk with cancer. Blood 2021, 137(23):3192–3200.

19. An SH, Kim IY: Can pretreatment platelet-to-lymphocyte and neutrophil-to-lymphocyte ratios predict long-term oncologic outcomes after preoperative chemoradiation followed by surgery for locally advanced rectal cancer? Annals of coloproctology 2022, 38(3):253.

20. Erdem GU, Topuz OV, Acar E, Kapagan T, Yetim E, Ozmen A, Gurocak S, Usul G, Yuksel S, Yardimci AH: Predicting complete response to neoadjuvant chemoradiotherapy in locally advanced rectal cancer: The role of baseline volumetric 18F-FDG PET/CT parameters and inflammatory markers. Revista Española de Medicina Nuclear e Imagen Molecular (English Edition) 2025:500113.

21. Karakaya S, Karadağ İ, Yılmaz ME, Öksüzoğlu ÖBÇ: High neutrophil-lymphocyte ratio, platelet-lymphocyte ratio and low lymphocyte levels are correlated with worse pathological complete response rates. Cureus 2022, 14(3).

22. Uyar GC, Başaran BN, Başkurt K, Yeşilbaş E, Özkan E, Yücel KB, Altınbaş M, Evrimler Ş, Öksüzoğlu ÖBÇ, Sütcüoğlu O: Predicting Pathologic Response in Locally Advanced Rectal Cancer Using Inflammatory, Nutritional, and Sarcopenia-Based Markers: A Regression and AI-Based Analysis (CINR-AI Study). Clinical Colorectal Cancer 2025.

23. Chiloiro G, Romano A, Mariani S, Macchia G, Giannarelli D, Caravatta L, Franco P, Boldrini L, Arcelli A, Bacigalupo A: Predictive and prognostic value of inflammatory markers in locally advanced rectal cancer (PILLAR)–A multicentric analysis by the Italian Association of Radiotherapy and Clinical Oncology (AIRO) Gastrointestinal Study Group. Clinical and translational radiation oncology 2023, 39:100579.

24. Xu YJ, Tao D, Qin SB, Xu XY, Yang KW, Xing ZX, Zhou JY, Jiao Y, Wang LL: Prediction of pathological complete response and prognosis in locally advanced rectal cancer. World J Gastrointest Oncol 2024, 16(6):2520–2530.

25. Yang W-N, Li X-M, Li C-F, Chen C, Feng Y, Dai N, Yang Y-X, Li M-X, Li C-X, Qian C-Y: Gustative Roussy Immune Score is a Predictor for Major Pathological Response in Rectal Cancer: A Result from the Preoperative Intraarterial Chemoembolization Combined with Radiotherapy (PCAR) Study. Cancer Investigation 2024, 42(6):527–537.

26. Page MJ, McKenzie JE, Bossuyt PM, Boutron I, Hoffmann TC, Mulrow CD, Shamseer L, Tetzlaff JM, Akl EA, Brennan SE: The PRISMA 2020 statement: an updated guideline for reporting systematic reviews. bmj 2021, 372.

27. Deutsch GB, Sathyanarayana SA, Gunabushanam V, Mishra N, Rubach E, Zemon H, Klein JD, Denoto G, 3rd: Robotic vs. laparoscopic colorectal surgery: an institutional experience. Surg Endosc 2012, 26(4):956–963.

28. Wells GA, Shea B, O’Connell D, Peterson J, Welch V, Losos M, Tugwell P: The Newcastle-Ottawa Scale (NOS) for assessing the quality of nonrandomised studies in meta-analyses. 2000.

29. Sterne JA, Savović J, Page MJ, Elbers RG, Blencowe NS, Boutron I, Cates CJ, Cheng H-Y, Corbett MS, Eldridge SM: RoB 2: a revised tool for assessing risk of bias in randomised trials. bmj 2019, 366.

30. Xun Y, Guo Q, Ren M, Liu Y, Sun Y, Wu S, Lan H, Zhang J, Liu H, Wang J: Characteristics of the sources, evaluation, and grading of the certainty of evidence in systematic reviews in public health: A methodological study. Frontiers in Public Health 2023, 11:998588.

31. Yang J, Deng Q, Chen Z, Chen Y, Fu Z: Body composition parameters combined with blood biomarkers and magnetic resonance imaging predict responses to neoadjuvant chemoradiotherapy in locally advanced rectal cancer. Frontiers in Oncology 2023, 13:1242193.

32. Zhou L, Cao G, Shi L, Fei C, Lao W: Risk factors of pathologic complete response for neoadjuvant chemoradiotherapy in locally advanced rectal cancer. Frontiers in Oncology 2025, 15:1483065.

33. Xiao B, Peng J, Zhang R, Xu J, Wang Y, Fang Y, Lin J, Pan Z, Wu X: Density of CD8+ lymphocytes in biopsy samples combined with the circulating lymphocyte ratio predicts pathologic complete response to chemoradiotherapy for rectal cancer. Cancer Management and Research 2017, 9(null):701–708.

34. Bozkaya Y, Özdemir NY, Erdem GU, Güner EK, Ürün Y, Demirci NS, Yazıcı O, Köstek O, Zengin N: Clinical predictive factors associated with pathologic complete response in locally advanced rectal cancer. Journal of Oncological Sciences 2018, 4(1):5–10.

35. Dudani S, Marginean H, Tang PA, Monzon JG, Raissouni S, Asmis TR, Goodwin RA, Gotfrit J, Cheung WY, Vickers MM: Neutrophil-to-lymphocyte and platelet-to-lymphocyte ratios as predictive and prognostic markers in patients with locally advanced rectal cancer treated with neoadjuvant chemoradiation. BMC cancer 2019, 19(1):664.

36. Lee IH, Hwang S, Lee SJ, Kang BW, Baek D, Kim HJ, Park SY, Park JS, Choi GS, Kim JC: Systemic inflammatory response after preoperative chemoradiotherapy can affect oncologic outcomes in locally advanced rectal cancer. Anticancer research 2017, 37(3):1459–1465.

37. Jeon BH, Shin US, Moon SM, Choi JI, Kim M-S, Kim KH, Sung S-J: Neutrophil to lymphocyte ratio: a predictive marker for treatment outcomes in patients with rectal cancer who underwent neoadjuvant chemoradiation followed by surgery. Annals of Coloproctology 2019, 35(2):100.

38. Karaca M, Salim E, Alemdar MS, Karaca ÖD, Arıcı MÖ: Prognostic significance of Neutrophil-to-Lymphocyte ratio in predicting complete pathological response in rectal cancer patients receiving neoadjuvant chemoradiotherapy. Medical Science Monitor: International Medical Journal of Experimental and Clinical Research 2024, 30:e943750–943751.

39. Cheong C, Shin JS, Suh KW: Prognostic value of changes in serum carcinoembryonic antigen levels for preoperative chemoradiotherapy response in locally advanced rectal cancer. World Journal of Gastroenterology 2020, 26(44):7022.

40. Jung SW, Park IJ, Oh SH, Yeom S-S, Lee JL, Yoon YS, Kim CW, Lim S-B, Lee JB, Yu CS: Association of immunologic markers from complete blood counts with the response to preoperative chemoradiotherapy and prognosis in locally advanced rectal cancer. Oncotarget 2017, 8(35):59757.

41. Kim S, Joo M, Yeo MK, Cho MJ, Kim JS, Jo EK, Kim JM: Small heterodimer partner as a predictor of neoadjuvant radiochemotherapy response and survival in patients with rectal cancer: A preliminary study. Oncol Lett 2021, 22(4):708.

42. Ren D-L, Li J, Yu H-C, Peng S-Y, Lin W-D, Wang X-L, Ghoorun RA, Luo Y-X: Nomograms for predicting pathological response to neoadjuvant treatments in patients with rectal cancer. World journal of gastroenterology 2019, 25(1):118.

43. Krauthamer M, Rouvinov K, Ariad S, Man S, Walfish S, Pinsk I, Sztarker I, Charkovsky T, Lavrenkov K: A study of inflammation-based predictors of tumor response to neoadjuvant chemoradiotherapy for locally advanced rectal cancer. Oncology 2013, 85(1):27–32.

44. Mehraj A, Baba AA, Khan B, Khan MA, Wani RA, Parray FQ, Chowdri NA: Predictors of pathological complete response following neoadjuvant chemoradiotherapy for rectal cancer. Journal of Cancer Research and Therapeutics 2022, 18(Suppl 2).

45. Soykut EDk, Kemal Y, Odabaşi E, Şahın N, Yağiz BKn, Yazicioğlu IrM, Arslan SA, Güney Y: Pre-Treatment and Post-Treatment Neutrophil-to-Lymphocyte Ratio Predict Pathological Tumor Response and Survival in Rectal Cancer Patients Treated with Neoadjuvant Chemoradiotherapy. Turkish Journal of Oncology 2022, 37(2).

46. Eraslan E, Adas YG, Yildiz F, Gulesen AI, Karacin C, Arslan UY: Systemic immune-inflammation index (SII) predicts pathological complete response to neoadjuvant chemoradiotherapy in locally advanced rectal cancer. J Coll Physicians Surg Pak 2021, 30(4):399–404.

47. Zhang F, Yu D, Yang J, Zhai M, Li L, Zhao L, Wang J, Zhang T, Lin Z: Pretreatment high cholesterol and low neutrophils predict complete pathological response after neoadjuvant short–course radiotherapy followed by chemotherapy and immunotherapy in locally advanced rectal cancer. Oncol Lett 2023, 26(1):319.

48. Zhou L, Cao G, Shi L, Fei C, Lao W: Risk factors of pathologic complete response for neoadjuvant chemoradiotherapy in locally advanced rectal cancer. Front Oncol 2025, 15:1483065.

49. Chiloiro G, Romano A, Mariani S, Macchia G, Giannarelli D, Caravatta L, Franco P, Boldrini L, Arcelli A, Bacigalupo A et al: Predictive and prognostic value of inflammatory markers in locally advanced rectal cancer (PILLAR) - A multicentric analysis by the Italian Association of Radiotherapy and Clinical Oncology (AIRO) Gastrointestinal Study Group. Clin Transl Radiat Oncol 2023, 39:100579.

50. Eraslan E, Adas YG, Yildiz F, Gulesen AI, Karacin C, Arslan UY: Systemic Immune-inflammation Index (SII) Predicts Pathological Complete Response to Neoadjuvant Chemoradiotherapy in Locally Advanced Rectal Cancer. J Coll Physicians Surg Pak 2021, 31(4):399–404.

51. Zhao Z, Xu H, Ma B, Dong C: Prognostic value of platelet to lymphocyte ratio (PLR) in breast cancer patients receiving neoadjuvant therapy: a systematic review and meta-analysis. Front Immunol 2025, 16:1658571.

52. Cullinane C, Creavin B, O’Leary DP, O’Sullivan MJ, Kelly L, Redmond HP, Corrigan MA: Can the Neutrophil to Lymphocyte Ratio Predict Complete Pathologic Response to Neoadjuvant Breast Cancer Treatment? A Systematic Review and Meta-analysis. Clin Breast Cancer 2020, 20(6):e675–e681.

53. Ma L, He J, Li P, Ma L, Wang H, Deng Y: Prognostic value of neutrophil to lymphocyte ratio in patients with esophagus cancer receiving neoadjuvant therapy: a systematic review and meta-analysis. Front Immunol 2025, 16:1615962.

54. Qu F, Luo Y, Peng Y, Yu H, Sun L, Liu S, Zeng X: Construction and validation of a prognostic nutritional index-based nomogram for predicting pathological complete response in breast cancer: a two-center study of 1,170 patients. Front Immunol 2023, 14:1335546.

55. Qu F, Li Z, Lai S, Zhong X, Fu X, Huang X, Li Q, Liu S, Li H: Construction and Validation of a Serum Albumin-to-Alkaline Phosphatase Ratio-Based Nomogram for Predicting Pathological Complete Response in Breast Cancer. Front Oncol 2021, 11:681905.

56. Hajian-Tilaki K: Receiver Operating Characteristic (ROC) Curve Analysis for Medical Diagnostic Test Evaluation. Caspian J Intern Med 2013, 4(2):627–635.

57. Mao Y, Pei Q, Fu Y, Liu H, Chen C, Li H, Gong G, Yin H, Pang P, Lin H et al: Pre-Treatment Computed Tomography Radiomics for Predicting the Response to Neoadjuvant Chemoradiation in Locally Advanced Rectal Cancer: A Retrospective Study. Front Oncol 2022, 12:850774.

58. Li A, He K, Guo D, Liu C, Wang D, Mu X, Yu J: Pretreatment blood biomarkers predict pathologic responses to neo-CRT in patients with locally advanced rectal cancer. Future Oncol 2019, 15(28):3233–3242.

59. Hou R, Wu X, Wang C, Fan H, Zhang Y, Wu H, Wang H, Ding J, Jiang H, Xu J: Tumor–associated neutrophils: Critical regulators in cancer progression and therapeutic resistance (Review). Int J Oncol 2025, 66(4).

60. Ozel I, Duerig I, Domnich M, Lang S, Pylaeva E, Jablonska J: The Good, the Bad, and the Ugly: Neutrophils, Angiogenesis, and Cancer. Cancers (Basel) 2022, 14(3).

61. Gordon-Weeks AN, Lim SY, Yuzhalin AE, Jones K, Markelc B, Kim KJ, Buzzelli JN, Fokas E, Cao Y, Smart S et al: Neutrophils promote hepatic metastasis growth through fibroblast growth factor 2-dependent angiogenesis in mice. Hepatology 2017, 65(6):1920–1935.

62. Zhou Y, Shen G, Zhou X, Li J: Therapeutic potential of tumor-associated neutrophils: dual role and phenotypic plasticity. Signal Transduct Target Ther 2025, 10(1):178.

63. Obeagu EI: N2 Neutrophils and Tumor Progression in Breast Cancer: Molecular Pathways and Implications. Breast Cancer (Dove Med Press) 2025, 17:639–651.

64. Mai J, Yang L, Chen Y, Zeng X, Xie H, Liu X: Prediction of CD8(+) T cell infiltration in the tumor microenvironment of HGSOC patients. Sci Rep 2025, 15(1):30518.

65. Liao K, Zhang X, Liu J, Teng F, He Y, Cheng J, Yang Q, Zhang W, Xie Y, Guo D et al: The role of platelets in the regulation of tumor growth and metastasis: the mechanisms and targeted therapy. MedComm (2020) 2023, 4(5):e350.

66. Guo Y, Cui W, Pei Y, Xu D: Platelets promote invasion and induce epithelial to mesenchymal transition in ovarian cancer cells by TGF-β signaling pathway. Gynecol Oncol 2019, 153(3):639–650.

67. Zhang L, Yuan Y, Deng Y, Wang L, Chen F: Platelet–circulating tumor cell crosstalk: A pivotal target in cancer diagnosis and therapy (Review). Oncol Rep 2026, 55(1).

68. Zhu J, Lian J, Xu B, Pang X, Ji S, Zhao Y, Lu H: Neoadjuvant immunotherapy for colorectal cancer: Right regimens, right patients, right directions? Front Immunol 2023, 14:1120684.

69. Farooq B, Li X, Xiao S, Liu X: Neoadjuvant therapy for pMMR/MSS locally advanced rectal cancer in the immunotherapy era: current landscape and future perspectives. Front Oncol 2025, 15:1719642.

