## Supplementary for "Immune-inflammatory Indices as Predictors of Complete Response to Neoadjuvant Chemoradiotherapy in Locally Advanced Rectal Cancer: A Systematic Review and Meta-Analysis of Observational Studies"

Table S1: Search Strategy

| Database | Search Strategy |
|---|---|
| **PubMed** | ((("Rectum"[Mesh]) OR ("Rectal"[Title/Abstract] OR "Rectum"[Title/Abstract] OR "rectal cancer"[Title/Abstract] OR "locally advanced rectal cancer"[Title/Abstract] OR "LARC"[Title/Abstract])) AND ("Neutrophil-to-Lymphocyte" OR "Neutrophil to Lymphocyte" OR "neutrophil/lymphocyte" OR "neutrophil-lymphocyte" OR "neutrophil lymphocyte" OR "Neutrophil-to-Lymphocytes" OR "Neutrophil to Lymphocytes" OR "neutrophil/lymphocytes" OR "neutrophil-lymphocytes" OR "neutrophil lymphocytes" OR "NLR" OR "platelet-to-Lymphocyte" OR "platelet to Lymphocyte" OR "platelet/lymphocyte" OR "platelet-lymphocyte" OR "platelet lymphocyte" OR "platelet-to-Lymphocytes" OR "platelet to Lymphocytes" OR "platelet/lymphocytes" OR "platelet-lymphocytes" OR "platelet lymphocytes" OR "PLR" OR "Systemic Immune-Inflammation" OR "Systemic Immune Inflammation" OR "SII")) AND (("Pathologic Complete Response"[Mesh]) OR ("complete response"[Title/Abstract] OR "complete pathologic response"[Title/Abstract] OR "pathologic response"[Title/Abstract] OR "pathologic complete response"[Title/Abstract] OR "pathological response"[Title/Abstract] OR "pCR"[Title/Abstract] OR "tumor regression grade"[Title/Abstract] OR "TRG"[Title/Abstract])) |
| **Embase** | ('rectal':ti,ab,kw OR 'rectum':ti,ab,kw OR 'rectal cancer':ti,ab,kw OR 'locally advanced rectal cancer':ti,ab,kw OR 'larc':ti,ab,kw) AND ('neutrophil-to-lymphocyte' OR 'neutrophil to lymphocyte' OR 'neutrophil/lymphocyte' OR 'neutrophil-lymphocyte' OR 'neutrophil lymphocyte' OR 'neutrophil-to-lymphocytes' OR 'neutrophil to lymphocytes' OR 'neutrophil/lymphocytes' OR 'neutrophil-lymphocytes' OR 'neutrophil lymphocytes' OR 'nlr' OR 'platelet-to-lymphocyte' OR 'platelet to lymphocyte' OR 'platelet/lymphocyte' OR 'platelet-lymphocyte' OR 'platelet lymphocyte' OR 'platelet-to-lymphocytes' OR 'platelet to lymphocytes' OR 'platelet/lymphocytes' OR 'platelet-lymphocytes' OR 'platelet lymphocytes' OR 'plr' OR 'systemic immune-inflammation' OR 'systemic immune inflammation' OR 'sii') AND ('complete response':ti,ab,kw OR 'complete pathologic response':ti,ab,kw OR 'pathologic response':ti,ab,kw OR 'pathologic complete response':ti,ab,kw OR 'pathological response':ti,ab,kw OR 'pcr':ti,ab,kw OR 'tumor regression grade':ti,ab,kw OR 'trg':ti,ab,kw) |
| **Scopus** | ( TITLE-ABS-KEY ( "Rectal" OR "Rectum" OR "rectal cancer" OR "locally advanced rectal cancer" OR "LARC" ) AND ALL ( "Neutrophil-to-Lymphocyte" OR "Neutrophil to Lymphocyte" OR "neutrophil/lymphocyte" OR "neutrophil-lymphocyte" OR "neutrophil lymphocyte" OR "Neutrophil-to-Lymphocytes" OR "Neutrophil to Lymphocytes" OR "neutrophil/lymphocytes" OR "neutrophil-lymphocytes" OR "neutrophil lymphocytes" OR "NLR" OR "platelet-to-Lymphocyte" OR "platelet to Lymphocyte" OR "platelet/lymphocyte" OR "platelet-lymphocyte" OR "platelet lymphocyte" OR "platelet-to-Lymphocytes" OR "platelet to Lymphocytes" OR "platelet/lymphocytes" OR "platelet-lymphocytes" OR "platelet lymphocytes" OR "PLR" OR "Systemic Immune-Inflammation" OR "Systemic Immune Inflammation" OR "SII" ) AND TITLE-ABS-KEY ( "complete response" OR "complete pathologic response" OR |

| | |
|---|---|
| | "pathologic response" OR "pathologic complete response" OR "pathological response" OR "pCR" OR "tumor regression grade" OR "TRG" ) ) |
| **Web of Science** | "Rectal" OR "Rectum" OR "rectal cancer" OR "locally advanced rectal cancer" OR "LARC" (Topic) and "Neutrophil-to-Lymphocyte" OR "Neutrophil to Lymphocyte" OR "neutrophil/lymphocyte" OR "neutrophil-lymphocyte" OR "neutrophil lymphocyte" OR "Neutrophil-to-Lymphocytes" OR "Neutrophil to Lymphocytes" OR "neutrophil/lymphocytes" OR "neutrophil-lymphocytes" OR "neutrophil lymphocytes" OR "NLR" OR "platelet-to-Lymphocyte" OR "platelet to Lymphocyte" OR "platelet/lymphocyte" OR "platelet-lymphocyte" OR "platelet lymphocyte" OR "platelet-to-Lymphocytes" OR "platelet to Lymphocytes" OR "platelet/lymphocytes" OR "platelet-lymphocytes" OR "platelet lymphocytes" OR "PLR" OR "Systemic Immune-Inflammation" OR "Systemic Immune Inflammation" OR "SII" (All Fields) and "complete response" OR "complete pathologic response" OR "pathologic response" OR "pathologic complete response" OR "pathological response" OR "pCR" OR "tumor regression grade" OR "TRG" (Topic) |
| **Cochrane Library** | ("Rectal" OR "Rectum" OR "rectal cancer" OR "locally advanced rectal cancer" OR "LARC"):ti,ab,kw AND ("Neutrophil-to-Lymphocyte" OR "Neutrophil to Lymphocyte" OR "neutrophil/lymphocyte" OR "neutrophil-lymphocyte" OR "neutrophil lymphocyte" OR "Neutrophil-to-Lymphocytes" OR "Neutrophil to Lymphocytes" OR "neutrophil/lymphocytes" OR "neutrophil-lymphocytes" OR "neutrophil lymphocytes" OR "NLR" OR "platelet-to-Lymphocyte" OR "platelet to Lymphocyte" OR "platelet/lymphocyte" OR "platelet-lymphocyte" OR "platelet lymphocyte" OR "platelet-to-Lymphocytes" OR "platelet to Lymphocytes" OR "platelet/lymphocytes" OR "platelet-lymphocytes" OR "platelet lymphocytes" OR "PLR" OR "Systemic Immune-Inflammation" OR "Systemic Immune Inflammation" OR "SII") AND ("complete response" OR "complete pathologic response" OR "pathologic response" OR "pathologic complete response" OR "pathological response" OR "pCR" OR "tumor regression grade" OR "TRG"):ti,ab,kw |

Table S2. Risk of Bias Assessment using NOS

| Study (year) | Design | Selection (0-4★) | Comparability (0-2★) | Outcome/Exposure (0-3★) | Overall Score (0-9★) | Quality of study (High/Moderate/Low) |
|---|---|---|---|---|---|---|
| Mehraj (2022) [40] | Retrospective cohort | 4/4★ | 0/2★ | 3/3★ | 7/9★ | High |
| Delikgöz Soykut (2022) [41] | Retrospective cohort | 4/4★ | 2/2★ | 3/3★ | 9/9★ | High |
| Eraslan (2021) [42] | Retrospective cohort | 4/4★ | 0/2★ | 3/3★ | 7/9★ | High |
| Yang (2023) [22] | Retrospective cohort | 4/4★ | 0/2★ | 3/3★ | 7/9★ | High |
| Zhou (2025) [24] | Retrospective cohort | 4/4★ | 2/2★ | 3/3★ | 9/9★ | High |
| Xiao (2017) [25] | Retrospective cohort | 4/4★ | 0/2★ | 3/3★ | 7/9★ | High |
| Xu (2024) [23] | Retrospective cohort | 4/4★ | 0/2★ | 3/3★ | 7/9★ | High |
| Zhang (2023) [45] | Retrospective cohort | 4/4★ | 0/2★ | 3/3★ | 7/9★ | High |
| Bozkaya (2018) [26] | Retrospective cohort | 4/4★ | 0/2★ | 3/3★ | 7/9★ | High |
| An & Kim (2022) [27] | Retrospective cohort | 4/4★ | 0/2★ | 3/3★ | 7/9★ | High |
| Dudani (2019) [29] | Retrospective cohort | 4/4★ | 1/2★ | 3/3★ | 8/9★ | High |
| Jung (2017) [35] | Retrospective cohort | 4/4★ | 0/2★ | 3/3★ | 7/9★ | High |

| | | | | | | |
|---|---|---|---|---|---|---|
| Karakaya (2022) [30] | Retrospective cohort | 4/4★ | 0/2★ | 3/3★ | 7/9★ | High |
| Kim (2021) [37] | Retrospective cohort | 4/4★ | 0/2★ | 3/3★ | 7/9★ | High |
| Krauthamer (2013) [39] | Retrospective cohort | 4/4★ | 1/2★ | 3/3★ | 8/9★ | High |
| Lee (2017) [31] | Retrospective cohort | 4/4★ | 1/2★ | 3/3★ | 8/9★ | High |
| Jeon (2019) [32] | Retrospective cohort | 4/4★ | 0/2★ | 3/3★ | 9/9★ | High |
| Erdem (2025) [43] | Retrospective cohort | 4/4★ | 0/2★ | 3/3★ | 9/9★ | High |
| Karaca (2024) [33] | Retrospective cohort | 4/4★ | 0/2★ | 3/3★ | 7/9★ | High |
| Chiloiroa (2023) [28] | Retrospective cohort | 4/4★ | 0/2★ | 3/3★ | 9/9★ | High |
| Cheong (2020) [34] | Retrospective cohort | 4/4★ | 0/2★ | 3/3★ | 7/9★ | High |
| Uyar (2025) [44] | Retrospective cohort | 4/4★ | 1/2★ | 3/3★ | 8/9★ | High |
| Yang (2024) [36] | Retrospective cohort | 4/4★ | 0/2★ | 3/3★ | 7/9★ | High |
| Ren (2019) [38] | Retrospective cohort | 4/4★ | 0/2★ | 3/3★ | 7/9★ | High |

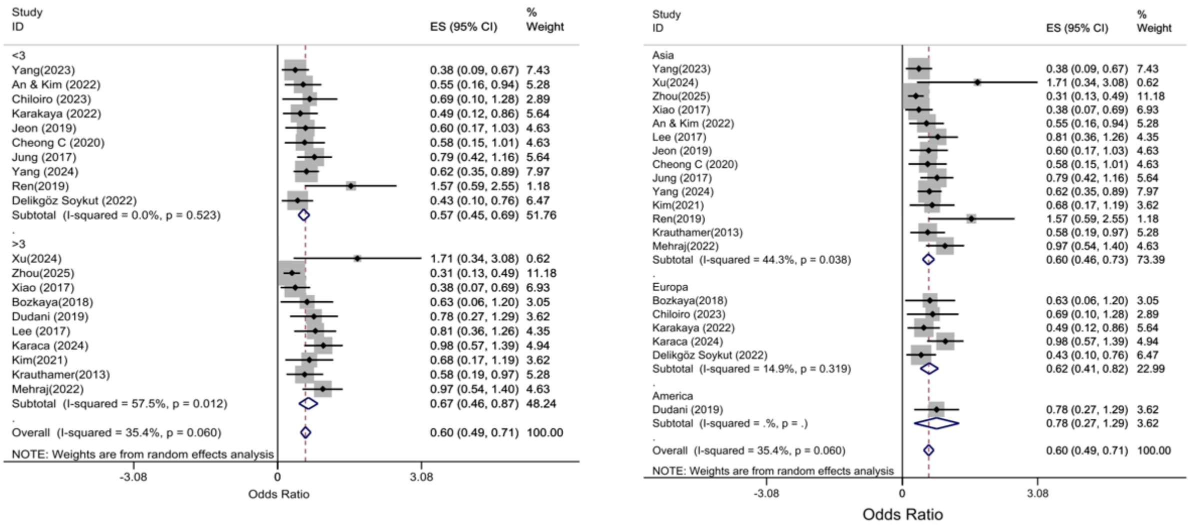


*Figure S1: Subgroup random-effects forest plots of pre-nCRT NLR vs pCR: Left panel, cut-off <3 vs ≥3; Right panel, region (Asia, Europe, America).*

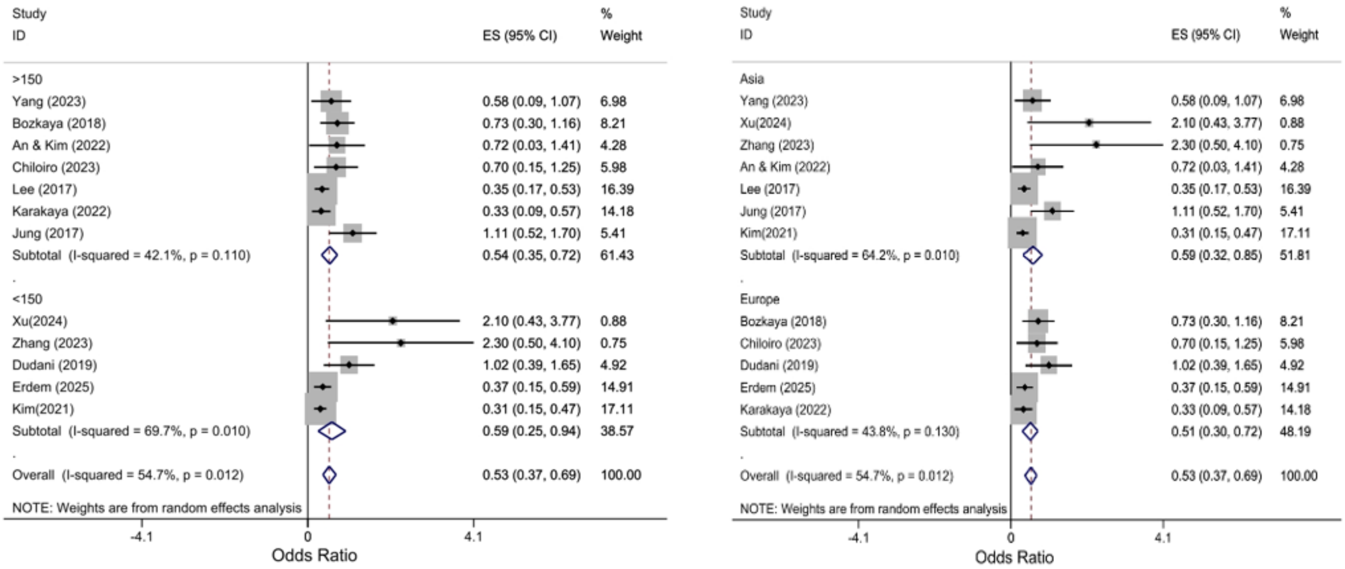


*Figure S2: Subgroup random-effects forest plots of pre-nCRT PLR vs pCR: Panel A, cut-off ≤150 vs >150; Panel B, region (Asia, Europe).*

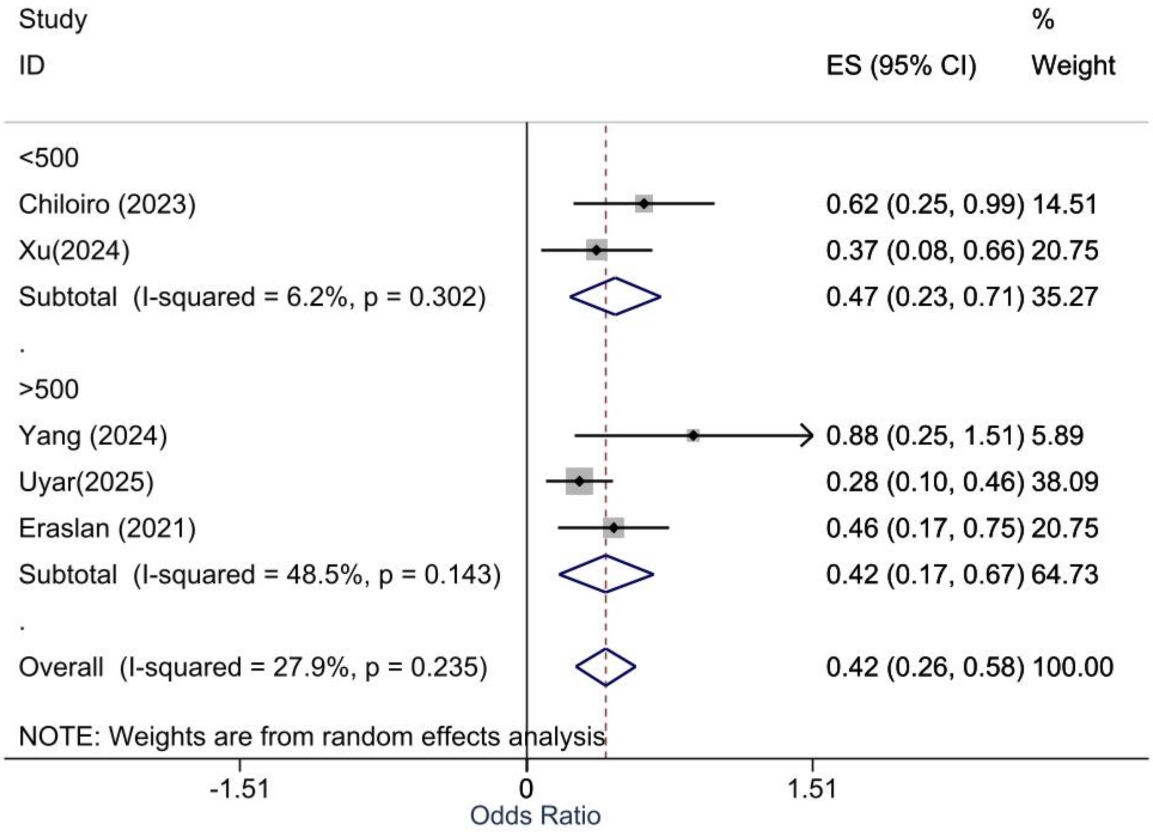


*Figure S3. Subgroup random-effects forest plot of the association between pre-nCRT PLR and pCR, stratified by PLR cut-off (≤500 vs >500).*

Table S3. Certainty of evidence (GRADE)

| Prognostic factor → Outcome | Studies (k) | Pooled effect (OR, 95% CI) | Overall certainty | Downgrade reasons |
|---|---|---|---|---|
| **Higher pre-nCRT NLR → lower pCR** | 20 | 0.60 (0.49–0.71) | Moderate | Consistent association in mostly high-quality cohorts with low–moderate heterogeneity; downgraded for suspected small-study effects. |
| **Higher pre-nCRT PLR → lower pCR** | 12 | 0.53 (0.37–0.69) | Low | Variable adjustment and cut-offs, moderate heterogeneity, and suspected small-study effects. |
| **Higher pre-nCRT SII → lower pCR** | 5 | 0.42 (0.26–0.58) | Low | Few studies with moderate heterogeneity, and potential small-study effects cannot be excluded. |

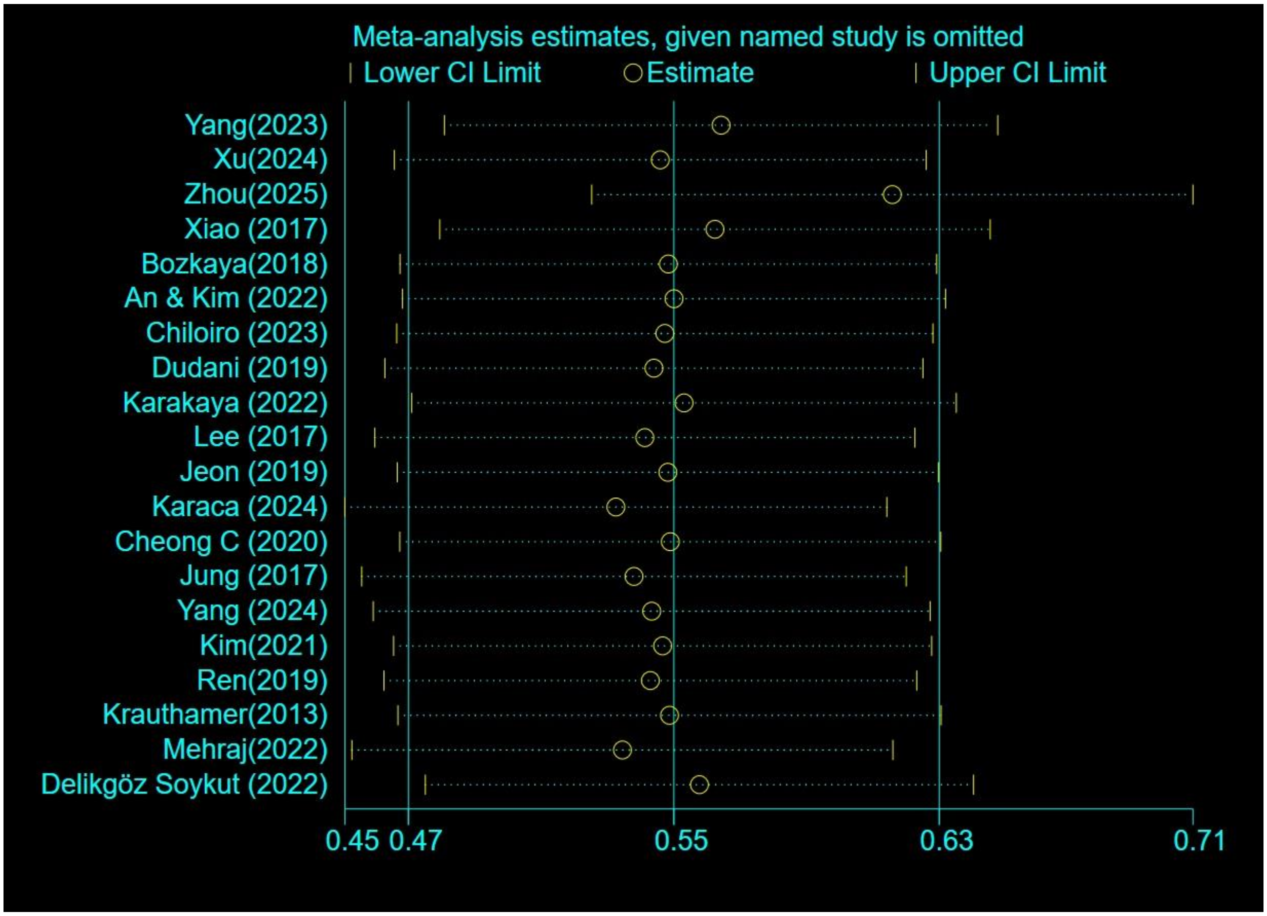


*Figure S4: Leave-one-out influence analysis for NLR: each point shows the pooled OR (with 95% CI) after omitting the named study.*

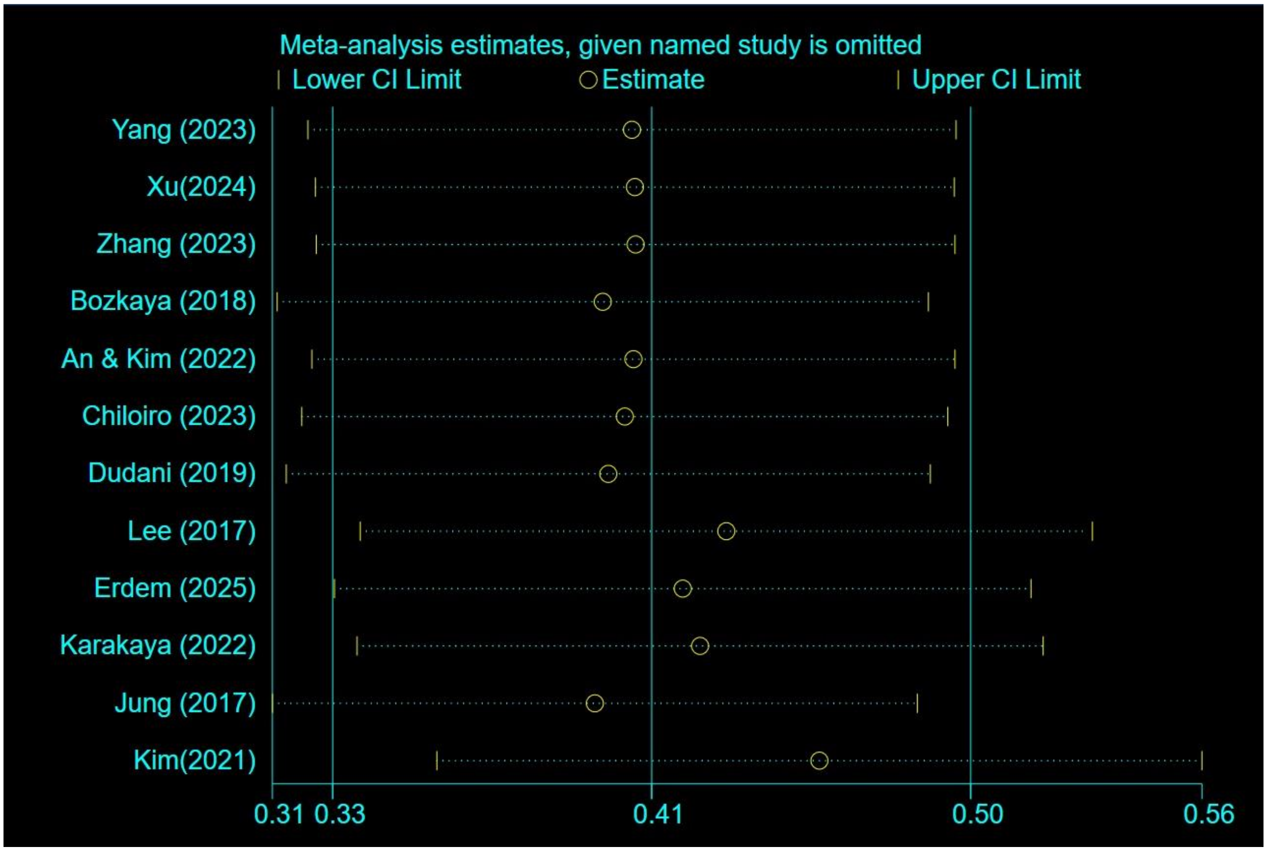


*Figure S5: Leave-one-out influence analysis for PLR: each point shows the pooled OR (with 95% CI) after omitting the named study.*

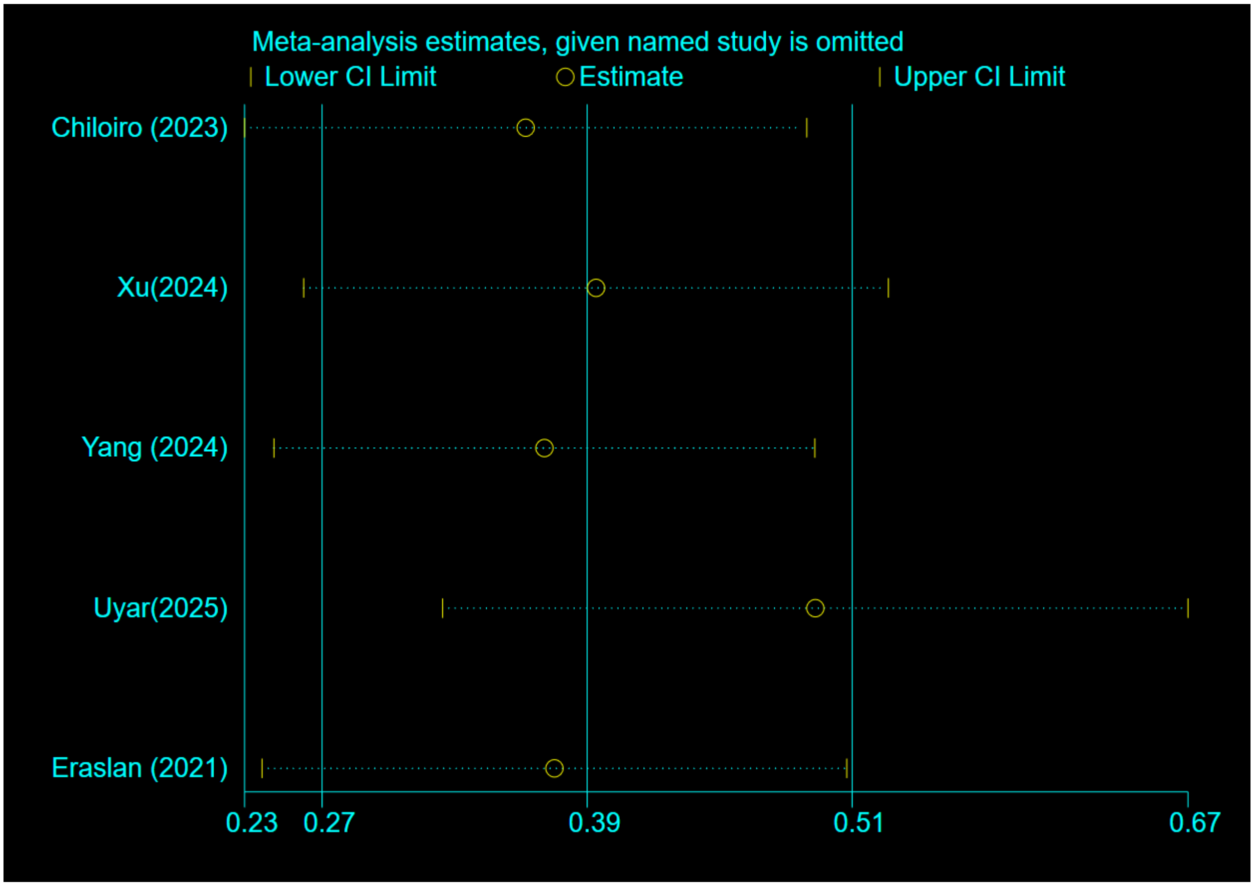


*Figure S6: Leave-one-out influence analysis for SII: each point shows the pooled OR (with 95% CI) after omitting the named study.*
